# Connecting Artificial Intelligence and Primary Care Challenges: Findings from a Multi-Stakeholder Collaborative Consultation

**DOI:** 10.1101/2021.09.21.21263906

**Authors:** Jacqueline K. Kueper, Amanda L. Terry, Ravninder Bahniwal, Leslie Meredith, Ron Beleno, Judith Belle Brown, Janet Dang, Daniel Leger, Scott McKay, Bridget L. Ryan, Merrick Zwarenstein, Daniel J. Lizotte

## Abstract

Despite widespread advancements in and envisioned uses for artificial intelligence (AI), few examples of successfully implemented AI innovations exist in primary care (PC) settings.

**Objectives:** To identify priority areas for AI and PC in Ontario, Canada.

**Methods:** A collaborative consultation event engaged multiple stakeholders in a nominal group technique process to generate, discuss, and rank ideas for how AI can support Ontario PC.

**Results:** The consultation process produced nine ranked priorities: 1) preventative care and risk profiling, 2) patient self-management of condition(s), 3) management and synthesis of information, 4) improved communication between PC and AI stakeholders, 5) data sharing and interoperability, 6-tie) clinical decision support, 6-tie) administrative staff support, 8) practitioner clerical and routine task support, and 9) increased mental health care capacity and support. Themes emerging from small group discussions about barriers, implementation issues, and resources needed to support the priorities included: equity and the digital divide; system capacity and culture; data availability and quality; legal and ethical issues; user-centered design; patient-centredness; and proper evaluation of AI-driven tool implementation.

**Discussion:** Findings provide guidance for future work on AI and PC. There are immediate opportunities to use existing resources to develop and test AI for priority areas at the patient, provider, and system level. For larger-scale, sustainable innovations, there is a need for longer-term projects that lay foundations around data and interdisciplinary work.

**Conclusion:** Study findings can be used to inform future research and development of AI for PC, and to guide resource planning and allocation.

**SUMMARY:** *What is already known?:*

– The field of artificial intelligence and primary care is underdeveloped.

*What does this paper add?:*

– An environmental scan without geographic location restriction identified 110 artificial intelligence-driven tools with potential relevance to primary care that existed around the time of the study.
– A multi-stakeholder consultation session identified nine priorities to guide future work on artificial intelligence and primary care in Ontario, Canada.
– Priorities for artificial intelligence and primary care include provider, patient, and system level uses as well as foundational areas related to data and interdisciplinary communication.

## INTRODUCTION

Advancements in artificial intelligence (AI) are leading to innovation in virtually every industry, including health care. In 2018, the WHO-UNICEF Global Primary Health Care Conference emphasized a need to effectively use current data and technology for innovations that will achieve better health care for individuals and populations [1]. Despite rich data sources and envisioned uses, the number of existing AI applications in primary care (PC) is smaller compared to other sectors [2–8]. To direct efforts and support more concrete progress towards development and use of AI for PC, interdisciplinary work that engages PC and AI stakeholders is needed to better understand how current and near-term AI capabilities align with existing PC challenges.

For study purposes, PC is defined as first contact and continuing care provided primarily by family physicians and nurse practitioners and excluding services provided solely by specialist care providers [9,10]. Ontario has a publicly funded health care system whereby PC is the entry point to the rest of the system. The structure of PC ranges from solo physician-based practices to large interdisciplinary teams, and different renumeration models exist such as fee-for-service and capitation-based models [11,12]. The field of AI began in the 1950s with a goal for computers to achieve human-like intelligence, e.g. making inferences or learning patterns from data, and has since expanded in the types of problems addressed and disciplines involved, such as psychology, philosophy, and linguistics [13]. The term “AI-driven tool” refers to technology that involves AI but may also have non-AI based functionality.

The objectives of this study were to identify current PC challenges that may be amenable to support using AI, discuss barriers and needs for successful development and implementation of AI to support those challenges, and identify priority areas for AI and PC in Ontario, Canada. We addressed these objectives by holding a multi-stakeholder collaborative consultation session in early 2021.

### Background: Environmental scans

As a preparation step for the stakeholder session, two brief environmental scans were conducted to better understand the landscape of PC needs and existing AI-driven tools. Environmental scans include identifying and summarizing information on a topic, often to support decision making [14].

The first scan explored PC challenges discussed in literature for high-income countries from 2010 through 2020, including those specific to the COVID-19 pandemic. Challenges were organized using a framework that divides PC into a structural domain for system-level considerations including practice context and organization, and a performance domain for service delivery and technical quality of clinical care [15]. Example challenges are presented in Table 1; detailed methods and results can be accessed at [16].

**Table 1.** Example primary care challenges discussed in literature from 2010 through 2020.

| Structural Domain | Performance Domain |
| --- | --- |
| General Primary Care Challenges |  |
| - Provider shortage | - Physician burnout |
| - Resource allocation does not meet current | - Need for improved coordination |
| demands<br>- Nurse practitioners not able to practice full scope of skills<br>- Inequitable access | - Difficulty of applying guidelines for patients with multimorbidity<br>- Need for improved relational continuity |
| **COVID-19 Specific Challenges** |  |
| - Lack of personal protective equipment<br>- Provider payment delays<br>- Nurse shortages in northern communities<br>- Need for early diagnosis and follow-up of high-risk patients | - Decreased use of primary care services<br>- Virtual care reduced human connection<br>- Care continuum challenges<br>- Patient backlogs |

The second scan identified 110 AI-driven tools with potential uses in PC. An estimated 87% (n=96) were in use at the time of identification. Table 2 presents tool characteristics and Figure 1 categorizes the tools by PC-related tasks they are intended to support; categories are based on a framework by EIT Health and McKinsey & Company for assessing impact of AI on health care [17]. Of note, these results focus on “ready-to-use” tools, but 36 active patents were also identified and a previous scoping review looked at the state of AI and PC research specifically [2]. Detailed methods and results are in Supplementary Material A.

**Table 2.**
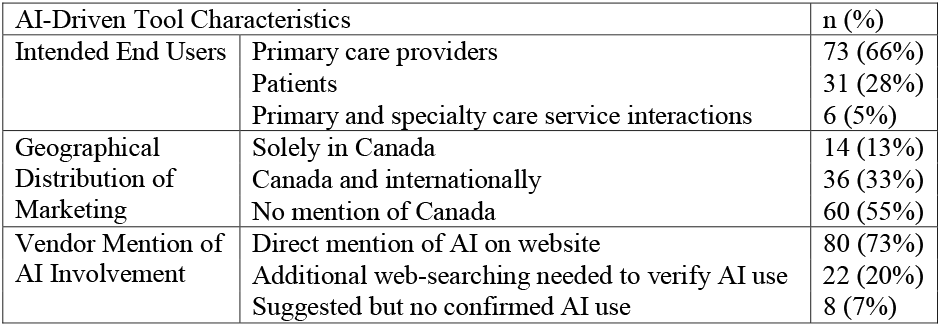
AI-Driven tool characteristics identified by environmental scan.

**Figure 1.**
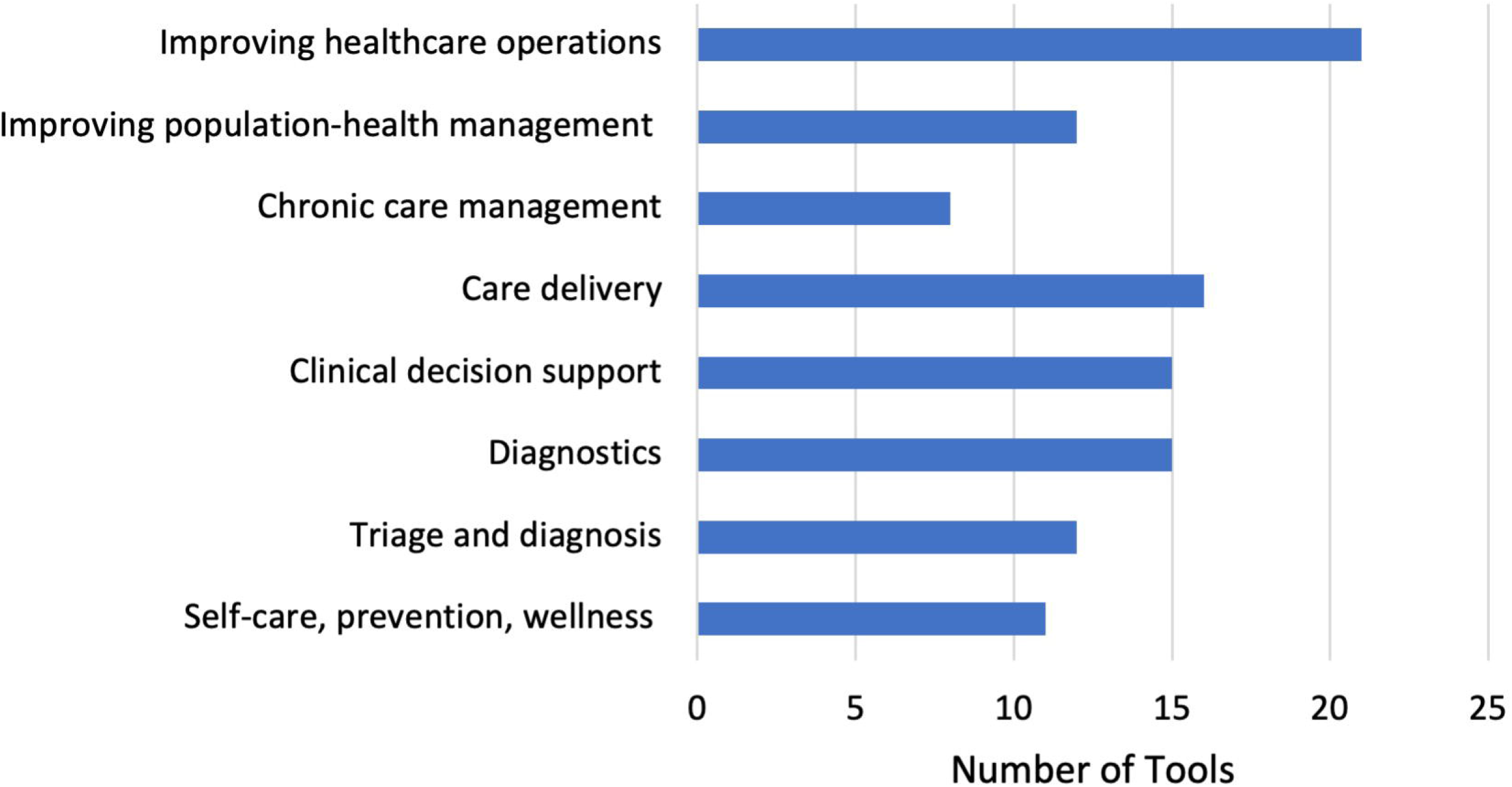
Application areas of AI-driven tools with potential relevance to primary care that existed around the time of the consultation session (details in Supplementary Material A).

## METHODS

### Participants

We invited patient (includes caregiver), provider, research, digital health, and decision-maker stakeholders with expertise or interest in AI and Ontario PC. Participants were recruited by e-mail individually utilizing the study investigators’ networks, with patients and caregivers recruited by the patient advisor co-investigator.

### Overview of agenda

The 4.5-hour multi-stakeholder consultation session was held on March 26, 2021 through Zoom. Primers introducing core AI and PC concepts were created and provided to participants in advance of the event. The morning agenda included a welcome and orientation, brief presentation on the environmental scans, and a keynote address by Dr. Winston Liaw on “The experience of utilizing AI in primary health care”. The afternoon included small and large group (all participants present) discussions, described below, with ranking activities according to a modified version of the nominal group technique [18,19]. Participants remained in the same small group throughout the event. Each group had a designated moderator, AI-knowledge resource person, and note taker; these roles could be fulfilled by one or more people. The AI-knowledge person was present to answer any technical questions and to ensure discussions did not stray from realistic current or near-term AI capabilities. Real-time, independent and anonymous ranking was done using Mentimeter [20]. Moderators and AI-knowledge people did not participate in ranking activities.

### Nominal group technique

The nominal group technique was developed to facilitate idea generation and discussion in a group setting for the purpose of arriving at a list of priorities [18,19]. It can be done within a day and allows for equal voting weights by all participants [18,19]. Steps included:

#### First small group discussion: ideating PC challenges

In each small group, participants were asked to independently write down answers to the question, “Thinking about PC, what issues do you think may be amenable to AI solutions?”. Participants were invited to share ideas in a roundtable format, each offering an idea with a brief discussion about why it is an issue for them, until no new unique ideas were generated. The group then worked together to collapse and clarify issues, if needed, to enter them into a Mentimeter poll. Finally, each participant was asked to “Rank based on what you feel are the top priorities for implementation in PC of issues that have a potential AI solution”. Each group ended Step 1 with up to 12 ranked issues.

#### First big group report back

In turn, each small group selected a member to share their Mentimeter chart and briefly describe the group’s top ranked challenges to the large group.

#### Second small group discussion: in-depth exploration of priorities

*S*mall groups re-convened for in-depth discussions of their top 2-3 ranked issues. Lower ranked items could be discussed with group consensus. Facilitated discussions encouraged participants to think about barriers, implementation issues or feasibility, and resources that would be needed to develop and/or implement AI to address the issues. Although discussions were facilitated on an item-by-item basis, there was overlap between groups and between items such that common themes applicable to all or most items emerged. Common themes that emerged are separated from item-specific points in the presentation of results.

#### Second big group report back

In turn, each small group had six minutes to report on their previous discussion and priorities to the large group.

#### Full group ranking activity

The priorities selected for discussion and reported on by each small group were merged into one list with similar items combined. Each participant ranked the entire list of items based on the criteria of, “Practicality, feasibility, and achievability within the next year or two”. Participants were shown ranking results in real-time and thanked for their participation.

After the stakeholder event, the research team reviewed the list of ranked priorities in combination with the small group discussion notes to construct a final list of priority areas. Wordings or descriptions were refined to increase clarity of presentation, maintaining the core content behind each ranked priority.

The project was approved by the Western Health Sciences REB (Project ID 116208).

## RESULTS

### Participants

There were 35 participants: 8 providers, 8 patient advisors, 4 decision-makers, 3 digital health stakeholders, and 12 researchers. Participants were divided into four pre-specified small groups, each with a range of stakeholder types.

### First small group discussion: Ideating primary care challenges

Complete lists of identified issues and rankings generated by each small group are in Supplementary Material B. A summary with similar items across groups collapsed includes:

- *Managing and/or consolidating information from different sources to facilitate identification of problems*.
- *Clinical decision support*.
- *Administrative Staff support*.
- *Patient self-management*.
- *Data sharing and interoperability between providers*.
- *Risk profiling and reminders for screening and preventive care*.
- *System coordination and referral centralization*.
- *Documentation and clerical duties*.
- *Patient triage in autumn (expected pandemic recovery phase) and help to manage and identify high-risk patients*.
- *Mental health care*.
- *Communication and adoption between AI and PC practitioners*.

### Second small group discussion: In-depth exploration of priorities

In discussing feasibility and necessary resources for successful development or implementation of AI to meet identified priorities, groups considered areas spanning from technical underpinnings of AI-driven tools to human and system-level factors. A synthesis of these discussions is below.

Groups emphasized **data availability and quality** as a foundation for successful AI development and application. Participants noted several different types and sources of potentially valuable data that exist, such as patient portals, text-based clinical notes, and structured electronic medical record (EMR) entries. Participants also noted challenges with the current state of EMR databases, and how it would be beneficial to work towards standardization and interoperability. They expressed concerns about learning from biased data, reconciling data from different sources (e.g., allergy reported in one database but not another), and the need for digital infrastructure and storage. While participants emphasized the need for long-term projects to develop high-quality PC databases, they also suggested small-scale AI projects with available data as a useful starting point.

Data considerations also emerged in conversations about **legal and ethical issues**. Challenges to be addressed included data ownership, including EMR vendors who can sell data, and data sharing with informed patient consent. Participants debated pros and cons of data sharing versus non-sharing as a standard setting, considering potential group benefit and personal privacy and control. Another unresolved barrier towards deployment of AI in PC settings is clarity around AI-driven tool certification to allow use in clinical settings.

In discussing the development of AI-driven tools, participants emphasized the need for **user-centred and ethical design** that includes input from patient, provider, and AI stakeholders, noting there may be heterogeneous preferences even within these stakeholder groups. Workflow considerations for tool design were also mentioned. In addition to design-oriented comments, participants discussed care-oriented needs, such as **patient-centredness** and fostering trust between patients, providers, and technology. Participants expressed that they did not want AI to block patient-provider relationships nor to act as an independent authority.

Once developed, there is a need for proper **evaluation of AI-driven tool implementation**. Participants envisioned starting with small-scale projects that take advantage of available data and working with “early adopters” or care teams at high-quality test sites to develop and test AI for PC settings. Participants encouraged thinking carefully about what meaningful evaluation will look like and what indicates success according to different stakeholders. Participants also mentioned a need for more research funding to conduct this type of research.

Conversations also considered the role of **system capacity and culture** and how organizational change will be at least as big a barrier to innovation as technical challenges are. Participants noted the need for intersectoral collaboration and for more integration and alignment between jurisdictions, especially for data regulations and linkage. The capacity to share information through communities, e.g., with patient social networks and social media, was also raised as a useful resource.

Finally, a central theme of caution around **equity and the digital divide** emerged throughout all discussions. Example concerns included access to required technology and consideration of populations who may have unique experiences or needs, such as older adults or those experiencing homelessness.

### Full group ranking activity: Final prioritized list

After combining similar items across small groups, there were nine AI and PC priorities to rank. Table 3 presents the final ranked list with extended descriptions based on notes taken in the second small group discussions. At a high-level, there are four areas that the priorities are intended to support: practitioners in a clinical setting (Priorities 1,6A,8); patients (priorities 2,9); system-level activities (priorities 6B,3); and foundational areas that would support the quality and efficiency of other priorities (priorities 4,5).

**Table 3.** Final list of priority areas for AI and PC identified and ranked in the multi-stakeholder collaborative consultation day.

| Rank | AI & PHC Priority | Extended descriptions from small group discussions |
| --- | --- | --- |
| 1 | Preventative care and risk profiling | **Overarching goal:** Support decisions in cases of uncertainty around screening and/or potential diagnoses, and to free up time during clinical consults.<br>**Example specific tasks/outcomes:**<br>• Screening reminders for patients at high risk of negative outcomes; reminders would be more personalized than general guidelines.<br>• Facilitate earlier diagnosis when potentially beneficial and mitigate unnecessary testing otherwise. |
| 2 | Patient self-management of condition(s) | **Overarching goal:** Support patient self-care or self-management of condition(s) with the possibility of sharing information between patients and providers.<br>**Example specific tasks/outcomes:**<br>• Vaccines, including COVID-19, or medication reminders.<br>• Health coaching and other resources to support goal achievement, including feedback on progress between clinical appointments.<br>• Scheduling and appointment reminders.<br>• Education about conditions and expectations. |
| 3 | Management and synthesis of information sources | **Overall goal:** Use, combine, and/or synthesize information from multiple sources to expand the scope of practice and improve equity and care access.<br>**Example specific tasks/outcomes:**<br>• Identify relevant information sources/content for different 1) users, 2) questions, and 3) tasks.<br>• Manage the overwhelming amount of information from multiple sources. |
| 4 | Improved communication between PC and AI stakeholders | **Overall goal:** Support communication between AI practitioners, PC practitioners, and patients to mitigate misunderstandings and poor application of techniques.<br>**Example specific tasks:**<br>• Establish a shared vocabulary/lexicon.<br>• Include PC, AI, and patient stakeholders on projects. |
| 5 | Data sharing and interoperability | **Overarching goal:** Improve data sharing and interoperability between providers and jurisdictions. |
|  | between providers | **Example specific tasks:**<br>• Establish data standards to enable interoperability.<br>• Establish data linkages between provinces and health systems.<br>• Highlight the potential for individuals to contribute through data sharing, similar to concepts used for organ donation and the greater good. |
| 6A (tie) | Clinical decision support | **Overarching goal:** Support care decisions during times of uncertainty and/or high demand by providing suggestions or support to clinicians.<br>**Example specific tasks:**<br>• Individualised recommendations for interventions at the individual or group level.<br>• Standardize and/or summarise information from EMRs so that both patients and clinicians have access and can track relevant data.<br>• Support patient triage decisions during times of high demand (e.g. COVID-19 pandemic recovery phase) and as clinicians adjust to different modes of delivery post-pandemic (e.g. continuation of virtual visit options). |
| 6B (tie) | Administrative staff support | **Overarching goal:** Support administrative staff and patient appointment preparation.<br>**Example specific tasks:**<br>• Scheduling.<br>• Patient triage and deciding appointment modality.<br>• Chat bot that interacts with patient to provide appointment reminders and gather logistical questions about appointments, e.g. preferred language and transportation needs. Gathered information can be communicated back to administrative staff to anticipate appointment needs in advance. |
| 8 | Practitioner clerical and routine task support | **Overarching goal:** Decrease the burden of routine tasks, such as documentation.<br>**Example specific tasks:**<br>• Centralized referral system between PC and specialty care.<br>• Automatic transcription/documentation to reduce note taking.<br>• Identify outstanding requisitions or tests needing follow-up.<br>• Group discussions note that referral centralization may be more important/appealing, but transcription seems more feasible in the short-term. |
| 9 | Increased mental health care capacity and support | **Overarching goal:** Support and/or increase the scope of mental health care from primary care settings.<br>**Example specific tasks:**<br>• Avatars and digital identity (non-live photos) to increase patient |
|  |  | comfort in seeking care.<br>• Decision support systems to mitigate ‘anchoring bias’ (move beyond initial information fixation to see/care for the whole person).<br>• Linking familial patients.<br>• Risk prediction and decision support.<br>• System level tools to avoid people falling through the cracks. |

## DISCUSSION

This study engaged patients and caregivers, providers, decision-makers, digital health, and research participants in a nominal group technique process to identify priority areas for AI and PC. Small group discussions identified barriers, implementation issues, and necessary resources for progress. The final list of nine priority areas included physician, patient, and system-level supports; and foundational areas that are necessary for the success of AI in other priority areas.

The consultation session revealed foundations that need to be improved to support progress in AI development and application, such as communication between PC and AI stakeholders, intersectoral collaboration, data standards and interoperability, and legal issues. In addition to longer-term foundational work, participants encouraged the initiation of AI projects that align with priorities and offered suggestions about conducting these projects in settings where the data and culture are in place to support the continuum of AI development through to careful evaluation and implementation. Core values to maintain throughout these processes included collaboration between diverse participants to maintain suitability of a candidate tool for practice settings, and to attend to equity concerns and patient-centredness.

Most of the ranked priorities from the consultation session include areas wherein AI may support PC by performing relevant functions or tasks, such as using AI to predict patients at high risk of poor health where early intervention is useful. It is noteworthy that despite this consultation session happening during the COVID-19 pandemic, with instructions that the pandemic should be considered in responses, only two COVID-19 specific priorities were identified in the first small group discussion and none remained in the final ranked list. It is also interesting that the environmental scan found 110 AI-driven tools that may be relevant to PC, yet amongst our participants selected for their engagement in AI and PC, few examples of AI-driven tools implemented in Ontario PC settings were discussed. Appraising specific tools to see if they are available in Ontario and whether they are suitable to meet priority areas as delineated in this study could be an avenue for future research.

The two “foundation-related” priorities (4 and 5) will support progress of AI for all areas of PC. Many AI applications for PC will rely on data generated by PC, which was the topic of Priority 5 and in small group discussions about data access, quality, and consent. Initiatives supporting use of Ontario PC data for research include Institute for Clinical Evaluative Sciences (ICES) [21] and the emerging PC Ontario Practice-Based Learning and Research (POPLAR) Network [22], as well as national databases such as those housed by the Canadian PC Sentinel Surveillance Network (CPCSSN) [23] and the Canadian Institute for Health Information [24]. Paprica et al. (2019) explored views of Ontario general public regarding the use of linked administrative health data held by ICES, finding that people are generally in favour of using these data for public benefit, assuming privacy and security; however, positive attitudes towards data use are more mixed or negative when there is private sector involvement [25]. These findings are congruent with our findings regarding the importance of data ownership and oversight for AI-driven tool development, which has substantial private sector involvement.

Data sharing and communication also was a priority in a study by Shaw et al. (2018) that used the nominal group technique to elicit priorities for virtual care related policy planning for Ontario PC [26]. Similar to AI, at the time of their consultation (before the COVID-19 pandemic) virtual care was considered a novel technology with potential benefit. Their recommendations included the need for a patient-centred focus and system- or social-level changes [26]. One suggestion for engaging patients was in outcome measure selection [26], which is relevant for AI applications as well which ties together the themes emerging from our study around patient centredness and the need for rigorous evaluation. Another relevant suggestion regarding virtual care implementation was the use of a sociotechnical model of care [26], which is also cited as important for AI to contribute to a learning health system framework whereby data are used in feedback loops to improve care [27]. The idea of a sociotechnical model aligns with themes from our small group discussions about the importance of system culture and communication between stakeholders. Previous work towards improving multidisciplinary collaborations includes guidelines produced by Saleh et al. (2020) for AI-clinical collaborations [28], co-design of a documentation assistant for PC consultations by Kocaballi et al. (2020) in Australia [29], and a “code to bedside” framework for quality improvement methods by Smith et al. (2021) in the United States [30].

Given the breadth and complexity of PC, there are many perceived opportunities for AI to be useful—focused efforts on tangible projects are needed for the field to mature. Our consultation session identified priority PC challenges, which AI is well suited to support given the current or near-term capabilities of AI and the Ontario PC context. Although the consultation sessions focused on Ontario, the environmental scans suggest there may be similarities in terms of AI-driven tools and PC needs in other jurisdictions. Other sectors may use our list of priorities as a starting point to refine based on their context. Together, the findings from our study can be used to guide future research and evaluation efforts, as well as to guide organizations and decision-makers in guiding the allocation of resources towards advancing AI for PC.

### Strengths and limitations

Strengths of the study, which is the first to identify priorities for AI and PC in Ontario, included bringing together multiple types of stakeholders to capture diverse perspectives. The study pre-work to both appraise the present context based on environmental scans and to provide primer documents provided foundational knowledge that supported strong engagement by all participants, regardless of prior AI knowledge. Limitations include representation mainly from academic communities as opposed to industry and private practice. The environmental scans were not limited in this way; therefore, help to balance these findings.

## CONCLUSION

A multi-stakeholder event was held to identify priority areas for AI and PC in Ontario, Canada, with additional findings related to barriers and resources that would be needed to fully realize the benefits of these technologies. Findings provide both specific topic areas to pursue as well as general guiding principles for future work on AI for PC. Together, these findings can serve as a platform for an action plan related to advancing AI for PC.

## Data Availability

Data related to ranked items are provided in the manuscript or supplementary material. Raw notes from small group discussion will not be made available.

## ACKNOWLEDGEMENTS

We would like to acknowledge and thank all participants of the stakeholder consultation session for their time and energy, with a special thanks to the patient and caregiver advisors for sharing their perspectives on artificial intelligence and primary care.

