## Supplementary Material A for "Connecting Artificial Intelligence and Primary Care Challenges: Findings from a Multi-Stakeholder Collaborative Consultation"

### Supplementary Material A: Artificial Intelligence Tool Environmental Scan

*Ravninder Bahniwal*

An environmental scan was conducted to identify existing artificial intelligence (AI) driven tools that might be relevant to primary care (PC). This scan included a systematic search of peer-reviewed and grey literature, websites of key organizations, registered patents, and a web-based search.

#### Methods

*Scope of Search*
For this report, PC will include all first-contact services by providers, while excluding any services provided solely by specialist care providers. AI-driven tools targeted for a) use by PC providers, b) use by patients in PC, and c) use for mediating the interactions between PC and other parts of the health care system were included. The tools themselves had to be a) described as AI, b) portrayed characteristics of AI, c) developed using AI subtypes, or d) existing tools that incorporate AI specific technology. Searches were initially conducted in Summer 2020 and update searches were performed in early 2021.

*Search Strategy*
The search strategy is presented in Table A-1. Once key organizations and databases were identified, their websites were hand-searched using the established search terms to identify potential AI-driven tools. Once potential tools were identified, a Google search was used to locate vendor websites and secondary websites describing the tool in more detail. If the tool used AI and was deemed appropriate for use in PC, details regarding the AI-driven tool such as description, location, and target users were inputted into an Excel spreadsheet. This process was followed for grey literature, published literature, and web searches. The number of published literature papers identified are in Table A-2. Patents were searched using Espacenet, an international patent database (European Patent Office, 2020) and the Canadian Intellectual Property Office (Government of Canada, 2020).

Table A-1: Development of the Search Strategy

| Steps | Details |
|---|---|
| Step 1: Develop key terms | *Primary Search Terms:* Artificial Intelligence (Tools), Primary (Health) Care<br><br>*Additional Key Words:* Family Medicine, Family Physicians, Machine Learning, Virtual Health Care, Electronic Medical Record, Healthcare, Intelligent Medicine |
| Step 2: Key Organizations | Hand search the following websites with primary search terms: Ontario MD, Athenahealth, Canada Health Infoway, Vector Institute for Artificial Intelligence, |

| | |
|---|---|
| | Kaiser Permanente, Institute of Electrical and Electronics Engineers (IEEE), Qianhai Institute of Innovative Research (QIIR), EPIC, eClinicalWorks, CERNER, AMS Healthcare, Nuance, MEDETECH, Allscripts, Binah.ai, Cloud DX, Big White Wall, JUNO, Sensely |
| Step 3: Grey Literature | Hand search the following websites with primary search terms: Health Quality Ontario, Canadian Agency for Drugs and Technologies in Health, Canadian Institute for Health Information (CIHI), Turning Research into Practice (TRIP), Open Grey, Canadian Medical Association Infobase, Canadian Institute for Advanced Research, The College of Family Physicians of Canada |
| Step 4: Web-based search | Google search with "Artificial Intelligence" AND "Primary Care", scan titles of results on pages 1 through 10 to identify tools |
| Step 5: Peer reviewed literature | Keyword Search Query of "Artificial Intelligence" AND "Primary Care" for Cochrane Library, PubMed, EMBASE, Cinahl, Web of Science, Scopus, IEEE Xplore, ACM Digital Library |
| Step 6: Patents | Search Espacenet and Canadian Intellectual Property Office with "Artificial Intelligence" AND "Primary Care", include only active patents with descriptions showing potential use in PC |

Table A-2: Peer Reviewed Literature Search Part of the AI-driven Tool Environmental Scan

| **Database** | **Total Number of Results** | **Final Number of Tools Included** |
|---|---|---|
| Cochrane Library | 0 | 0 |
| Pub Med | 154 | 5 |
| EMBASE | 104 | 0 |
| Cinahl | 17 | 0 |
| Web of Science | 4 | 0 |
| SCOPUS | 108 | 0 |
| IEEE Xplore | 41 | 0 |
| ACM Digital Library | 118 | 2 |

*Data Extraction and Organization*

An Excel spreadsheet was used to keep a record of the findings throughout the search, including details of the company, description of the AI-driven tool, whether it explicitly mentioned relevance to PC or if this was referred, whether AI was mentioned alongside the tool, geographical location where the tool was being marketed, target end user, and category of usage. The use categories were taken based on a framework developed by EIT Health and McKinsey & Company (EIT Health, 2020). Descriptions of the categories are in Table A-3. For patents believed to have applications in PC, the title and description of the patent, applicant name and location was noted in the spreadsheet.

Table A-3: Descriptions of Categories Assigned to Identified Tools

| Category | Description of Tools |
|---|---|
| Patient Level | |
| Self-care, prevention and wellness | Aimed at supporting people to live healthier lives, monitoring and tracking devices (ex. vital signs), provide personalized guidance to individuals |
| Triage and early diagnosis | Symptom checkers that help triage patients and provide guidance if additional healthcare resources are required, helpful when there are long waiting times |
| Diagnostics | Help with diagnosis, such as when further clinical work is needed to determine underlying reasons for symptoms, often focus on specific, well-defined tasks |
| Clinical Decision Support | Retrieve relevant medical information for each patient and present it in a structured way, help physicians decide on best treatment option, determine patients at high risk of deterioration and complications, provide guidance for early intervention |
| Care Delivery | Often Natural Language Processing-based solutions that support practitioners during their direct interaction with patients, able to take notes, retrieve information from medical records, includes monitoring and treatment devices used in delivery of care |
| Chronic Care Management | Help patients manage their chronic diseases and continue care without needing hospitalization |
| System Level | |
| Improving population health management | Analyze large datasets that may uncover correlations between factors or identify early risk factors that can trigger early intervention and prevention at a system level, helps determine what to prioritize |
| Improving healthcare operations | Decrease time spent on routine, low-value administrative tasks to increase direct time with patients, helps with tasks occurring in the background of patient care such as scheduling and capacity management |

Framework developed by EIT Health, 2020.

**Results**

A total of 127 existing AI-driven tools with potential relevance to PC and an additional 36 active patents were identified. A complete list of identified tools is available upon request. Given the challenging nature of the search, this may not be an exhaustive list, but it does give an indication of the range of tools that are either currently influencing PC or showing promise with their development. A minority of tools stated a direct use in PC while most tools were targeted towards a variety of health care settings of which PC may be one. Example tools that specify a direct interest in PC are in Table A-4.

Table A-4: Descriptions of Companies and Their Tools That Stated a Direct Use in PC

| Company | AI-Driven Tool |
|---|---|
| Kaiser Permanente<br>*American integrated managed care consortium* | HIV prediction tool uses machine learning algorithm to predict who would become infected with HIV during a three-year period. This tool can be incorporated into electronic health records to alert PC providers to speak with patients most likely to benefit from discussions about PrEP. |
| University of Nottingham<br>*public research University* | An AI based clinical decision support software that scans patients' routine medical data and predicts which of them would have heart attacks or strokes within 10 years. It is intended for use by PC doctors while the patient is in front of them during a routine appointment or in a systematic screen of the entire list. |
| Brain FX<br>*medical technology manufacturer* | Brain FX assessment tools create a brief brain health profile of strengths and weaknesses in 10 to 15 minutes with automated reporting and advanced real-time analytics for baseline comparison or cohort analysis to determine next steps. |
| Saykara<br>*AI software start-up* | Kara is the first fully ambient intelligent virtual assistant. Kara interprets conversations with patients so you can enter the exam room, treat the patient and be done charting. Kara can create SOAP notes, place orders, write referrals, and complete pre-charting by reaching out to the patient ahead of time. |
| Suki<br>*AI Software company* | Suki is an AI-powered, voice-enabled digital assistant for doctors that generates accurate notes and gets smarter with each interaction. Doctors finish their notes an average of 76% faster with Suki. |

| | |
|---|---|
| Tyto Care<br>*telehealth start-up* | TytoHome is a remote examination tool and telehealth platform that connects consumers to clinicians to enable a comprehensive medical examination (ears, nose, throat, lungs, heart, temperature). |
| Cambridge Cognition<br>*cognitive assessment software provider* | CANTAB Mobile is a digital memory assessment tool for PC that can triage memory complaints in clinic. |
| Skin Analytics<br>*research led company committed to helping more people to survive skin cancer* | DERM AI is a machine learning tool that identifies malignant skin lesions with high levels of sensitivity and specificity. |
| Eyenuk Inc.<br>*global medical technology company* | EyeArt AI Eye Screening System makes in-clinic, real-time diabetic retinopathy screening possible for PC practices, diabetes centres, and optometric offices by allowing physicians to quickly and accurately identify referable DR patients during a diabetic patient's regular exam. |

*AI documentation.* Before classifying a tool as having AI capabilities, vendor websites were hand-searched for indications of AI use. Use of AI was directly mentioned on the vendor websites for 96 (76%) tools, whereas 23 (18%) required an additional web-search to determine the use of AI technologies. For 8 (6%) tools, the use of AI was unclear both on vendor websites and with a web-based search. Instead, AI use was inferred based on a deeper examination of website content. It should be noted that websites were not consistent when discussing the technology used by their tools. A trend can be seen with AI and software technology companies offering the most detail on the intricacies of their technology, whereas larger companies known for their EMR systems did not include many technical details. It should be noted that it is difficult to find complete information on the tools such as use cases, settings in which they should be implemented and how they were developed. While websites often included a page to input contact information so the vendor could directly contact a potential customer regarding a demo or further details, key information about the tools was not always publicly available. This finding suggests that end users could experience difficulties finding comprehensive information about the AI tools that exist, leading to potential skepticism and lack of trust in implementing new technologies.

*Locations*

When seeking the locations for the AI-driven tools, 17 (13%) were found to exist solely in Canada, 40 (31%) were found both in Canada and internationally, and 70 (55%) did not mention any use in Canada. It was noted that overall, 24 (19%) of the vendor companies are based in the United States region known as “Silicon Valley”, a global centre for innovative technology companies. These companies were also observed to have the most comprehensive information on their tools. Equally, four vendor companies based in Israel provided a similar level of information on their products as the Silicon Valley-based companies. This observation aligns with the Israeli government initiative to maximize usefulness for AI and encourage partnerships between foreign and domestic business, Israeli start-ups and health organizations (EIT Health, 2020). These findings align with technology sectors of countries such as US, UK, China and Israel building their health data repositories and investing in AI (Strome, 2020). Canada currently spends billions to purchase electronic health record systems from US vendors, and in the future will have to import costly AI technologies if it does not develop and scale AI applications within its borders (Strome, 2020).

*Target end user.* Of the total 127 identified tools, 81 (64%) were targeted towards PC providers, ranging from virtual assistants and risk assessment tools to devices aiding in diagnosis. There were 38 (30%) intended for patient use and can help with monitoring vital signs, checking symptoms, and managing chronic diseases. Of the tools intended for patient use, 12 (32%) are capable of remotely screening patients and directing them to appropriate healthcare services. If proven effective, these tools have the potential to reduce clinical workloads in PC while creating more time for providers to address patients with urgent needs. Another 8 (6%) tools were focused on the interaction between PC and specialist care services and can provide significant benefit in regions lacking specialty care access. For example, tools that detect diabetic retinopathy, malignant skin lesions and atrial fibrillation can be used when a referral is not possible or when long wait times create a delay before care can be provided by a specialist. The 36 active patents were not organized further as their target end-users in PC were often unclear and were used only to recognize the degree of innovation in progress.

*Categories of Tools*

As seen in Figure 1 of the manuscript, an overwhelming number of tools are targeted towards improving healthcare operations, aligning with current discussions on documentation practices contributing to increased physician burnout and reduced time spent with patients. Many tools in use focus on reducing time spent in administrative tasks such as record keeping and appointment scheduling. Speech recognition technologies and virtual assistants for notetaking are being utilized to enable PCP to focus exclusively on patient interactions. For patients accessing PC, there is an emphasis on empowering individuals to take control of their health through triage and diagnosis tools available on mobile applications. There are also several devices which track information such as vital signs and other wellness data to provide personalized guidance. Example tools for each category are in Figure A-1.

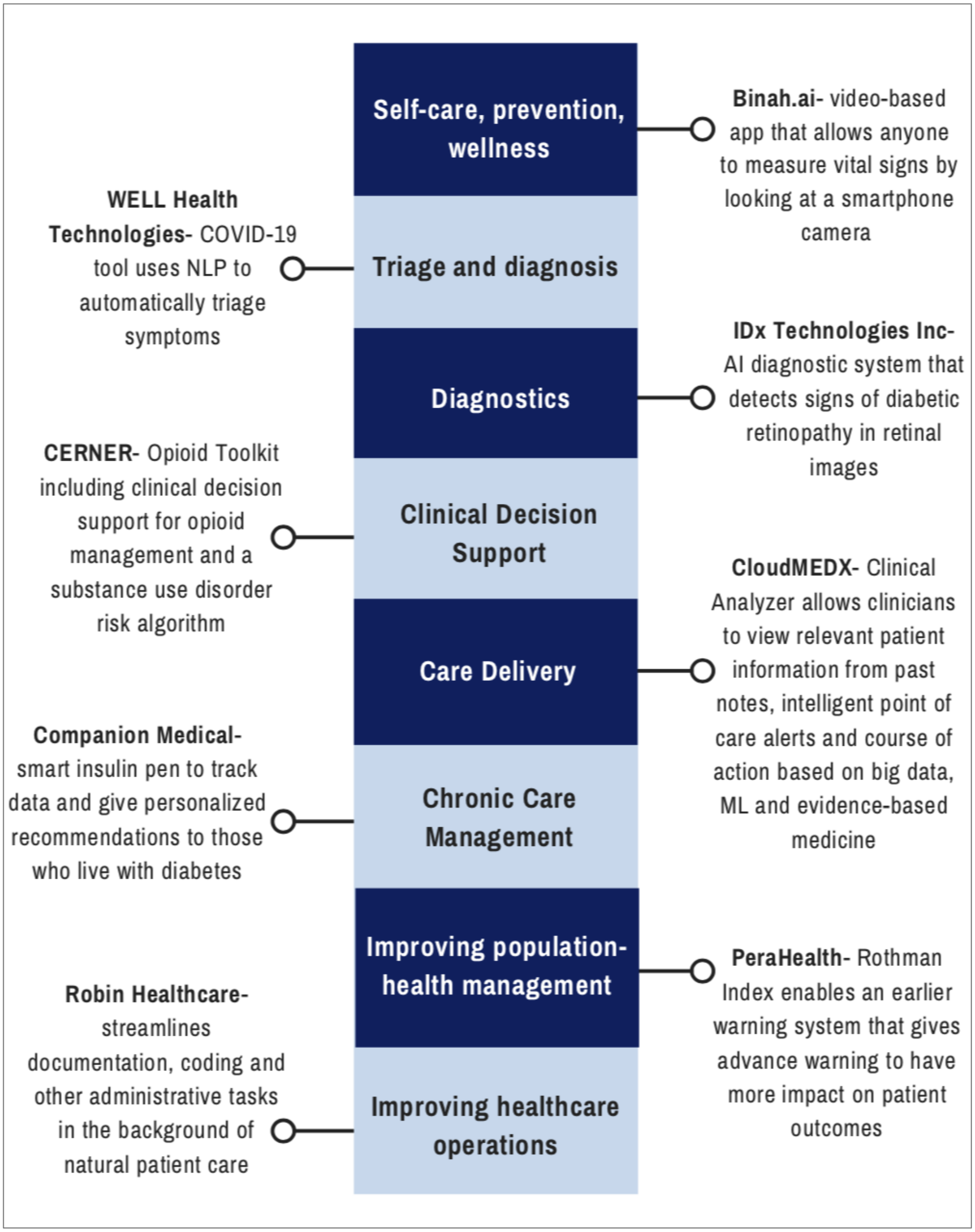


Figure A-1: Examples of AI-Driven Tool for Each Category

Based on the available data about the AI-driven tools, it is estimated that approximately 106 (83%) of the examined tools are currently in use somewhere in the world, with active patents confirming a future of further innovation. While scanning peer-reviewed literature, there were many articles focused on algorithms in development, along with discussion on the future of clinical decision support systems for specific diseases. Together with active patents, this observation may offer insight into the tools in development. Although many tools have EMR integration capabilities, there are still several tools developed independently by start-ups and information technology companies. In order to maximize their use in PC, it is important to connect PC providers and patients with the tools capable of solving current challenges in PC.
