## Supplementary Material B for "Connecting Artificial Intelligence and Primary Care Challenges: Findings from a Multi-Stakeholder Collaborative Consultation"

**Supplementary Material B: Small Group Discussion 1 Identified Areas**

Items were generated during the first small group discussion. Bolded items were discussed in the second small group discussion and the presented wording is from the small group discussion notes; other items are worded according to group's Mentimeter ranking chart from the first big group report back and thus may be more abbreviated.

<u>Small Group 1</u>

1. **Consolidating information from different sources to facilitate identification of problems**
2. **Clinical decision support**
3. **Administrative staff support, e.g. decision making around what type of care (virtual/in person), triaging, scheduling**
4. **Patient self-management**
5. Automated translation of language-literacy level
6. Portal medication view, visit/calendar view
7. Prescription renewal –patient driven synthesis of vast information – clinical decision support
8. Appropriate intervention as soon as possible – aggregate data
9. Bridge use of technology with patient comfort – equity concept
10. Vaccine alert, administrative function
11. Triage – link patient with right person

<u>Small Group 2</u>

1. **Patient self-management of disease is challenging, especially with multimorbidity. Patients have challenges in self-management of multimorbidity**
2. **Manage the firehose of information –** new medical knowledge is created, data is in charts, how can we use that newly created knowledge combined with what is in the chart to update care/suggest tests/develop or diagnose – expanding scope of practice and improving equity/access
3. **Data sharing and interoperability between/among providers**
4. AI could support changes to models of care – as a catalyst/lubricant that connects big data to people to improve the health care system
5. Primary care providers have to deal with many mundane workflow issues that could be made less painful
6. Information does not always flow effectively from providers to patient and caregiver

<u>Small Group 3</u>

1. **Risk profiling and reminders for screening and preventative care – use AI to personalize and go beyond general guidelines**
2. **System coordination and referral centralization**
3. **Documentation and clerical duties**
4. Patient level clinical decision making
5. Treatment plan prioritization
6. Population health management – identify unseen patterns and proactive reach out

7. Data collection
8. **Patient triage in the fall – identify high risk patients and how to manage**
9. Medical education resource amalgamation
10. Population health management – patient profiling/uncertainty
11. New ways of communication
12. Flexibility and human contact

Small Group 4

1. **Preventative care**
2. **Mental health care**
3. **Communication and adoption between AI and PC practitioners**
4. Reducing paperwork / clinical overhead
5. Handling patient contributed data
6. Assisting prioritization of care
7. Bedside manner assistance
8. Improving accuracy
9. Reducing inefficiencies
10. Communication challenges
11. Solving abandoned patients
